# Rapid prototyping of models for COVID-19 outbreak detection in workplaces

**DOI:** 10.1101/2023.02.05.23285483

**Authors:** Isobel Abell, Cameron Zachreson, Eamon Conway, Nicholas Geard, Jodie McVernon, Thomas Waring, Christopher Baker

## Abstract

Early case detection is critical to preventing onward transmission of COVID-19 by enabling prompt isolation of index infections, and identification and quarantining of contacts. Timeliness and completeness of ascertainment depend on the surveillance strategy employed. We use rapid prototype modelling to quickly investigate the effectiveness of testing strategies, to aid decision making. Models are developed with a focus on providing relevant results to policy makers, and these models are continually updated and improved as new questions are posed. The implementation of testing strategies in high risk settings in Australia was supported using models to explore the effects of test frequency and sensitivity on outbreak detection. An exponential growth model is firstly used to demonstrate how outbreak detection changes with varying growth rate, test frequency and sensitivity. From this model we see that low sensitivity tests can be compensated for by high frequency testing. This model is then updated to an Agent Based Model, which was used to test the robustness of the results from the exponential model, and to extend it to include intermittent workplace scheduling. These models help our fundamental understanding of disease detectability through routine surveillance in workplaces and evaluate the impact of testing strategies and workplace characteristics on the effectiveness of surveillance. This analysis highlights the risks of particular work patterns while also identifying key testing strategies to best improve outbreak detection in high risk workplaces.

## Introduction

Accurate and timely case detection is a key pillar of COVID-19 monitoring and management, particularly for countries that have aimed for zero community prevalence of SARS-CoV-2. Australia is an example of a country that had a ‘strong suppression’ policy until vaccines were widely distributed throughout 2021. Until that time, COVID-19 spread was prevented by proactive management of borders, active case finding and follow up with strict isolation and quarantine requirements for cases and contacts, and through liberal access to PCR testing in both high risk settings and the general community. Arriving international travellers posed the greatest risk of imported infection, leading to imposition of mandatory hotel quarantine arrangements on the 28^th^ of March that were maintained through to late 2021 [1]. Repeated SARS-CoV-2 incursions to the community, despite stringent arrivals procedures, prompted refinement of the design of testing strategies for travellers and workers in quarantine settings. Testing was one of a suite of layered infection prevention and control interventions to minimise the risk of community outbreaks.

Accuracy and completeness of case detection is influenced by multiple factors including: the dynamics of host-pathogen interaction, test performance (sensitivity and specificity), and test frequency. PCR tests are seen as the ‘gold standard’ in testing practices, although their performance varies over the course of the infection [2], with many individuals remaining PCR-positive after they are no longer infectious. For the purposes of outbreak detection, PCR tests suffer from their slow turnaround time (typically days) [2]. On the other hand, Rapid Antigen Tests (RATs) typically have a turnaround time of 15 minutes. However, they have lower sensitivity and specificity than PCR tests [3]. Similar to PCR tests, test sensitivity varies depending on the stage of infection, and whether infection is symptomatic or asymptomatic [3, 4]. Existing work suggests that these issues can be overcome by increasing the frequency of testing, implying that RATs can still be practically useful despite their limitations [5, 6].

In early 2021, Australia was seeking to improve testing strategies in workplaces, with the primary objective of detecting new outbreaks quickly. However, the differences in performance characteristics between different tests made it challenging to develop a robust workplace testing strategy. In addition to test performance, there were also questions about how the emergence of variants of concern, with differing characteristics such as transmissibility or severity, would affect outbreak detection. In this work, we describe the methods and results applied to rapid prototyping of testing strategies that were used to guide the Australian Government response of COVID-19 in 2021.

Rapid prototype modelling is a model development approach that aims to provide rapid insights while laying the foundations for more detailed modelling [7]. Models developed should be simple yet still convey complexities and subtleties of a problem to decision makers [8]. Problem identification at each step drives prototyping, and as questions are revised and improved, models are updated to reflect new scenarios [7, 9]. Rapid response modelling has been crucial in developing policy for COVID-19, allowing key hypotheses and assumptions to be tested in real-time as public health policy is implemented [10].

To provide timely advice on the principles underlying a robust workplace testing policy we took a rapid prototypic approach. We estimated the sensitivity of alternative surveillance strategies using models of differing granularity. The first is the ‘exponential model’, which we developed to give timely insight into testing efficacy, prompted by the emergence of the Alpha variant, which was both more transmissible and more pathogenic than antecedent viruses [11]. The model is defined by exponential growth of disease prevalence in the workplace, and allows us to quickly understand the interactions between test sensitivity, frequency and growth rate (*R*_eff_) on outbreak detection. The second model developed is an agent based model (ABM) which allows us to investigate the interaction between scheduled testing frequency and shift work patterns in determining the overall sensitivity of the surveillance system. The ABM builds on assumptions made by the rapidly developed exponential model to probe and update its results to answer new, emerging questions for an updated situation (i.e. intermittent work scheduling). These two models were developed as tools to provide answers to specific questions posed by policy makers about designing effective testing practices in workplaces.

We present our two models and results in the sequence they were developed, starting with the exponential model, which we use to explore the effects of growth rate (*R*_eff_), test sensitivity and test frequency on the probability of detecting outbreaks in workplaces. We then consider the Agent Based Model (ABM), using it to probe the results of the exponential model and also to explore the impact of more complex work schedules on the probability of outbreak detection. For both the exponential model and the ABM, we define an outbreak occurring when at least one employee is infected. An outbreak is detected when a positive case is identified, either through testing or symptom onset.

## Exponential model

Our initial model for the probability of detecting a COVID-19 outbreak in a workplace assumes exponential growth of active cases. In the exponential model disease prevalence on day *i* + 1 (denoted *P*_*i*+1_) is given by:

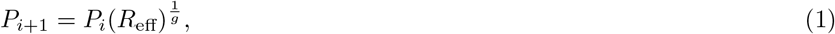

where *P*_*i*_ is the disease prevalence on day *i, g* is the generation interval and *R*_eff_ is the effective reproduction number. We assume a generation interval of 4.7 days, and that an outbreak begins on the first day with one active infection [12]. With these assumptions, we calculate the expected number of active cases through time under different values of *R*_eff_.

Using our model of prevalence, we then calculate the probability of detecting at least one case within a week under different testing strategies. We vary testing strategies by considering different test sensitivities and test frequency, i.e. the number of days per week testing occurs. In our results we consider scenarios where testing occurs once per week, three times per week and daily. We assume that on days when testing occurs, the entire workforce is tested. This testing framework is suitable for high importation risk, such as quarantine hotels.

The one-week detection probability is defined as the probability a truly infected individual returns a positive test within a week of the initial infection. Let *P*_*i*_ be the prevalence on testing day *i*, and *s* the test sensitivity. We assume tests have 100% specificity. The probability an outbreak is *not* detected on day *i* (given everyone is tested) is:

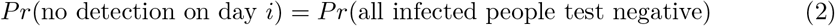

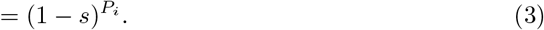

Let *T* be the set of testing days in a given week, i.e. days where everyone is tested. The probability of detection in a week is therefore given by:

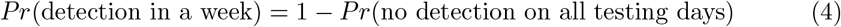

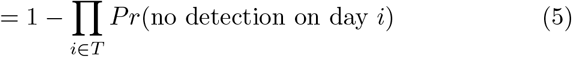

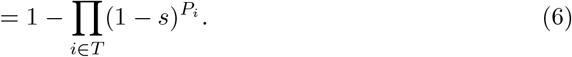

### Exponential model results

The probability of detection within a week increases with both test sensitivity and *R*_eff_ (Figure 1a). With testing occurring once per week, there is a large difference between whether there is low (0.65) or high (0.95) test sensitivity. However, the difference in the probability of outbreak detection between low and high sensitivity decreases with increasing *R*_eff_. As *R*_eff_ increases the outbreak spreads faster, meaning more infected people are tested within a week, increasing the likelihood that at least one is detected.

**Figure 1:**
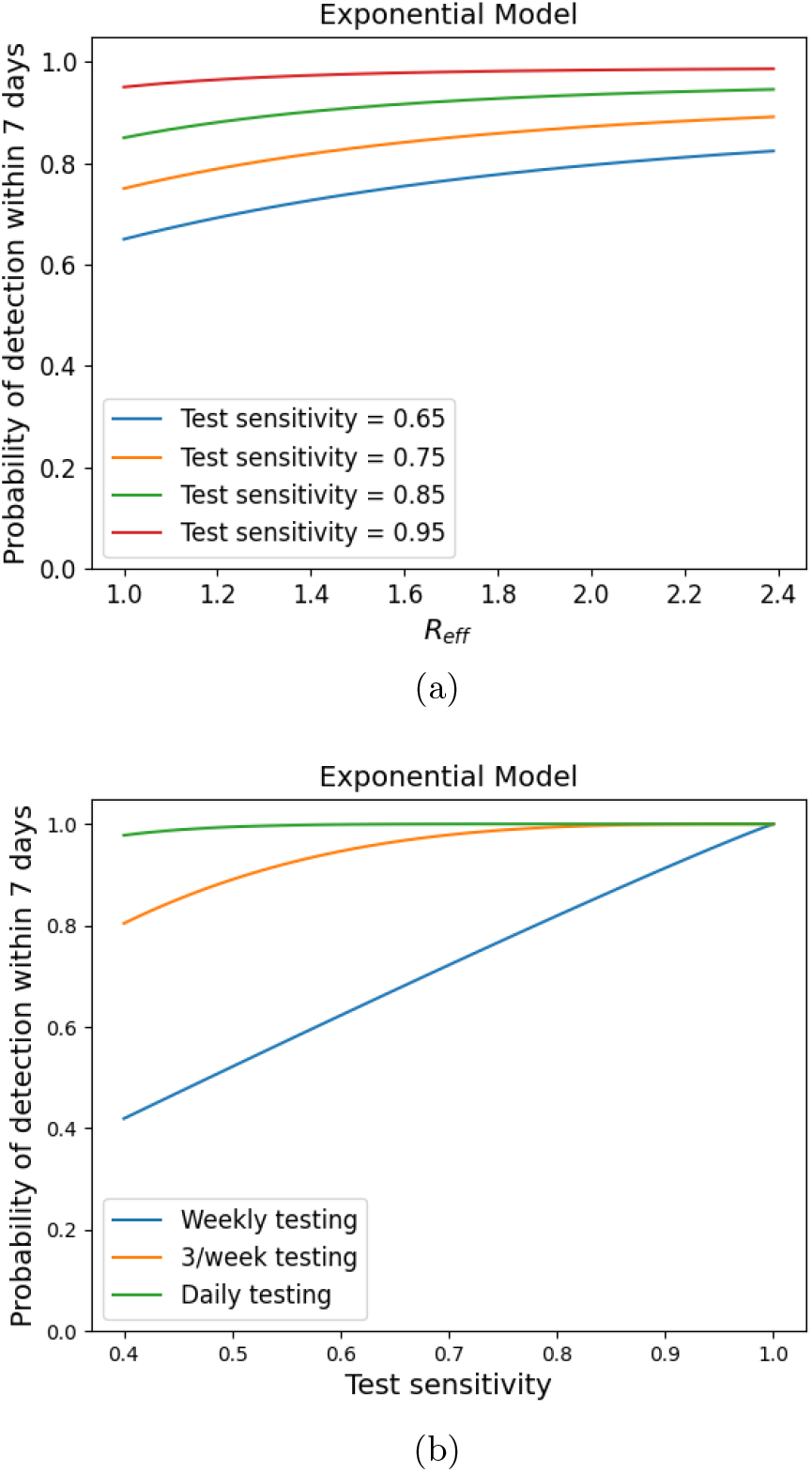
Probability of detection within a week using the exponential model as we vary growth rate (*R*_eff_) and test sensitivity (Figure 1a), and test sensitivity and testing schedule (Figure 1b). We assume a generation interval of 4.7 days and a workplace size of 50 people for both Figures 1a and 1b. For Figure 1a we assume testing occurs only once per week and for Figure 1b we assume a conservative growth rate of *R*_eff_ = 1.1.

Increasing test sensitivity and frequency both increase the probability of outbreak detection within a week (Figure 1b). Most notably, daily testing results in a high probability of detection within a week (*>* 95%), for all test sensitivities. This demonstrates that low-sensitivity tests, such as RATs, are still useful for outbreak detection — their shortcomings can be overcome by more frequent testing.

### Agent based model

We develop an agent based model (ABM) to represent additional complexities of the quarantine setting not captured in the exponential model. The ABM incorporates further complexity and allows us to ask more detailed questions about workplace testing. We start by setting up the ABM using the same set of assumptions as the exponential model. In replicating the exponential model results using the ABM, we can be confident that the ABM generalises the earlier results.

Each agent in our model follows an *susceptible*–*exposed* –*infectious*–*recovered* disease progression. Agents begin each simulation *susceptible*, and once infected become *exposed*. An exposed agent is neither infectious nor detectable. Exposed agents will transition to an *incubating* phase, where they become both infectious and detectable by testing. In the incubating phase, agents are either *symptomatic* or *asymptomatic*. Both infection states are detectable by testing, but symptomatic infection is also detected at the moment of symptom onset. When their infection ends, agents become *recovered*, and immune to reinfection. Further modelling details, including parameter values, can be found in the supplementary material.

For each simulation, outbreaks are seeded via a single infection in the workplace. As for the exponential model, we assume there are no cases imported into the workplace. An outbreak is detected when a positive test result is returned (assuming 100% specificity as for the exponential model) or symptom onset occurs. Under the ABM assumptions, we assume that two thirds of infectious people are symptomatic. To reproduce results from the exponential model, we assume all infected agents are asymptomatic, i.e. there will be no outbreak detection via symptom onset. For each model simulation, we estimate the probability of detecting at least one case within a week over 5000 simulation instances, each of which is subject to stochastic variation. We discard simulations instances in which all infected individuals recover before any are detected.

We define test frequency by specifying the number of testing days per week. As in the exponential model, on days when testing occurs, everyone attending the workplace is tested.

To extend the exponential model results, we consider the effects of intermittent work schedules on outbreak detection. We define these work schedules by specifying the proportion of the workplace working 1, 3, 5 or 7 days a week. Employees are then randomly assigned work days in accordance with the number of days they are scheduled to work. We assume people are only tested at work, but outbreaks may be detected through symptom onset at any time, regardless of whether the unwell individual is present in the workplace.

#### Agent Based Model Results

In this section, we present results from the ABM. We start by reproducing results from the exponential model to see how the ABM aligns with previous results. We then update our assumptions to consider the impact of intermittent workplace attendance on the probability of outbreak detection within a week of the introduction of the virus.

##### Comparing the exponential and agent based models

In line with the process of rapid prototyping, Figure 2 compares the exponential model behaviour to that of the ABM. Under identical sets of assumptions, the ABM results closely follow those of the exponential model (Figure 2a), although the ABM produces slightly more optimistic estimates of detection probability. However, when we change assumptions of the ABM (Figure 2b), the results begin to diverge. Under the new assumptions, the results from the ABM produce much higher probabilities of outbreak detection than the exponential model. This is explained by the additional mode of detection, by symptom onset. The additional assumptions we use here cannot be built into the exponential model due to its simplicity, so the development of the ABM allows us to explore the impact of these infection and testing characteristics on outbreak detection.

**Figure 2:**
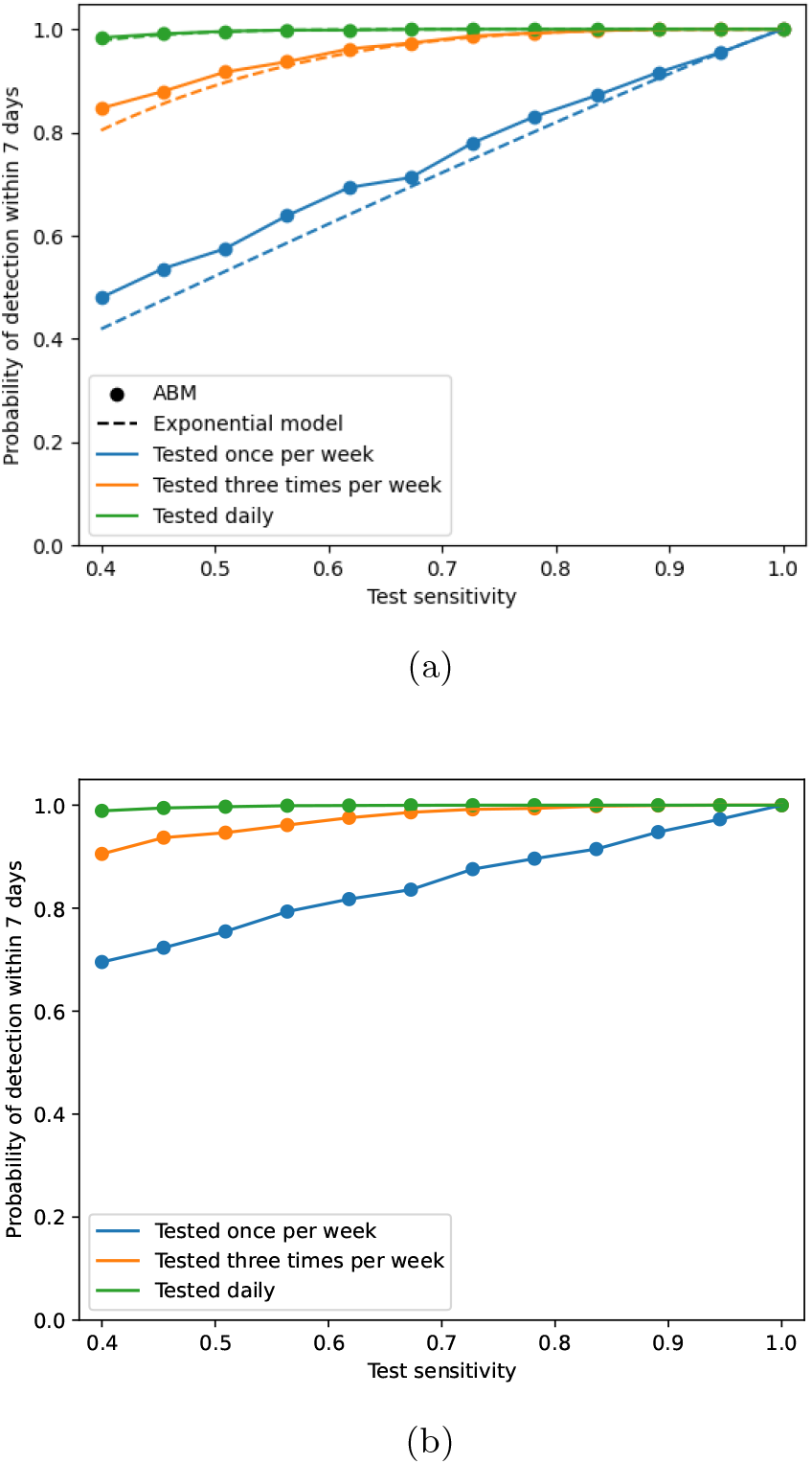
Probability of detection within a week as calculated by the exponential and agent based models as we vary growth rate, test sensitivity and test frequency. Figure 2a compares the ABM results to the exponential model under the same assumptions, i.e. no latent infection period, no asymptomatic infection and no detection via symptom onset. Figure 2b shows the ABM results under different assumptions to the exponential model, i.e. a latent infection period (1 day), asymptomatic infection and detection via symptom onset.

##### Intermittent workplace attendance

A strength of the ABM is that it can be used to explore the implications of more complex patterns of workplace attendance. We introduce an intermittent work schedule, where some proportion of workers work 1, 3, 5 and 7 days a week — though we assume that disease progression happens independently of this schedule. Analogously to the testing assumptions of the exponential model, we assume that when testing occurs, everyone in the workplace on that day is tested. We consider the following intermittent work schedules defined by the proportion of the workforce working 1, 3, 5 or 7 days a week:

1 100% 7 days/week,

2 100% 5 days/week,

3 60% 5 days/week, 40% 3 days/week,

4 60% 5 days/week, 30% 3 days/week, 10% 1 day/week.

As observed in the exponential model, increasing test frequency and sensitivity increases the probability of detecting an outbreak (Figure 3). In a similar way, the detection probability is higher when employees work more frequently. Notably, even for a sparse work schedule, low test sensitivity can be compensated for with higher testing frequency. If we increase test frequency, employees are more likely to be tested in a given week as they are more likely to be at work on a testing day. This compounds the benefits of frequent testing observed in the exponential model.

**Figure 3:**
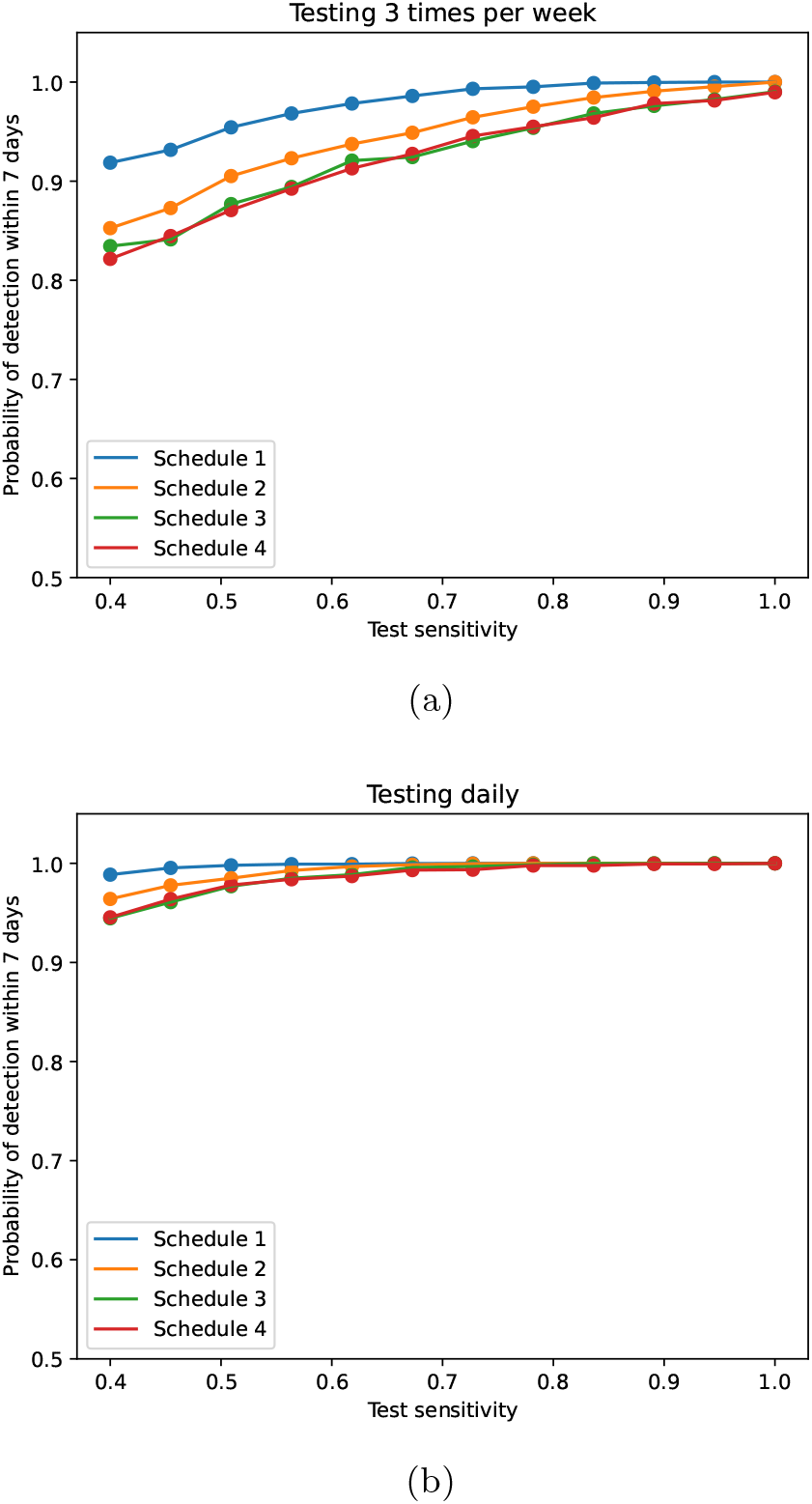
Probability of detecting an outbreak within a week under various intermittent working schedules as we vary test sensitivity and frequency for (a) testing three times per week and (b) testing daily. See text for details of testing schedules.

The ABM includes the assumption that outbreaks can be detected by symptom onset, which imposes an upper bound on the time to detection. Since disease progression occurs outside the workplace, outbreaks can be detected even when the infected workers are at home. This bound increases the seven-day detection probability compared to the exponential model.

## Discussion

Pandemic policies need to be adaptable in the face of emerging epidemic intelligence, including changes in circulating pathogen characteristics, host-pathogen interactions and diagnostic modalities [13]. By reassessing and updating our prototype models to examine new questions posed, we can aid decision makers by supporting evidence–based decisions as new scenarios arise. The development of the exponential model and the ABM demonstrates how a rapid prototyping approach is useful to for informing disease-management policy. While simple by design, the exponential model was quickly able to show that lower sensitivity tests can be useful when combined with high frequency testing, and that variants with higher *R*_0_ may be more readily detected in outbreak settings than less transmissible strains.

Answering these early questions naturally led to more nuanced questions, and we developed our second model in response to this. The ABM explores how the interaction between shift patterns and routine surveillance testing frequency determines the effective testing rate across the workforce. In our model, a high symptomatic proportion (*>* 60%) and perfect compliance with testing requirements mitigated identified risks associated with gaps in surveillance due to non-work days. These assumptions were valid in context of an unvaccinated population, circulation of the Alpha variant and strict public health orders mandating testing requirements. Assumptions must be updated in light of population and pathogen characteristics. For example in the case of COVID-19, by late 2022 a much lower symptomatic proportion would be expected given high levels of population vaccine coverage in Australia and emergence of the less pathogenic Omicron variant.

To answer questions around testing strategies in workplaces, our modelling aimed to model the decision problem at hand rather than simulate an outbreak as realistically as possible. Therefore, it is important to note that when designing models using a rapid prototyping approach, we cannot guarantee that the model will be suitable *outside* the scope of the question it is designed to answer. That is, it is not necessary that the models we use are good models for the system, but that they are instead suitable for the given questions. The exponential model is not considered a “good” model for understanding the spread of disease in workplaces, but, when focusing on a short time frame, it is useful for questions about outbreak detection. When new questions arise requiring nuance in the initial growth rate or longer term analysis, we need to turn to different models. In our context, the limitations of the exponential model led us to develop the agent based model. Despite the exponential model limitations, our understanding of the model allowed us to be confident in our results, and exploring a simple model gives us general insights about the decision problem. It also provides a solid foundation for further exploration with more complex models.

While Australia’s COVID-19 risk environment is continually evolving, both our modelling and the rapid prototyping framework can still provide useful insights. In contrast to the COVID-19 landscape in early 2021, Australia now has widespread COVID-19 transmission, meaning there are fewer workplaces where we actively seek to detect new outbreaks [14]. However, there remain workplaces, for example aged care facilities, where there is a high chance of severe outcomes from COVID-19, and so we would still seek to detect new outbreaks quickly to put in place mitigation measures. Furthermore, our models are fairly general, so our results are not specific to COVID-19. The exponential model assumes that we are aiming to detect something that is increasing in prevalence through time, and the only COVID-19 specific assumption is about the generation interval. Similarly, the ABM is quite general and these models could be readily adapted for other pathogens.

Our study shows the utility of taking a rapid prototyping approach to model development in epidemiology, starting by developing simple models and then building in additional complexity. Rapid prototyping has been used effectively for environmental management as part of Structured Decision Making (SDM) approaches [7, 9], but has not been used formally in epidemiology. Like ecological fields, epidemiology is well-suited for rapid prototyping due to its range of well-known simple models, e.g. SIR type models. In our example of workplace outbreak detection, the exponential model results provide a pessimistic estimate for the probability of outbreak detection. With updated information, the ABM provides a more realistic estimate of the probability of outbreak detection. Here, rapid prototyping allows us to provide a quick, conservative estimate to policy makers which can then be updated as more information becomes available.

The COVID-19 pandemic has highlighted the importance of model-generated evidence in decision making. With a short time-frame in which to answer questions, and a rapidly changing set of circumstances, flexible models which can be updated to new questions have an important role. The rapid prototyping process we describe is well suited to informing policy in a quickly evolving situation. While the importance of gaining quick insights for policy is clear, an additional benefit is that rapid prototyping models provide direction for development of more complex models. Simple models can provide useful insights to inform strategic thinking, and more detailed models are able to incorporate important real world complexities to refine tactics for surveillance and response.

## Data Availability

The code used to generate all results in this manuscript is available online

https://github.com/iabell/rapid_prototyping_covid_19

## Author’s contributions

IA led study design in collaboration with CB, EC, NG, JM and CZ. Preliminary modelling done by CB, EC, NG, JM, CZ and extended by IA. Manuscript written by IA and TW, and edited by all authors.

## Acknowledgements

This work was supported by the Australian Government Department of Health and Ageing Office of Health Protection.

## Supplementary material

For further details on parameter values and testing schedules, please see the supplementary material.

## Code availability

The code used to generate all results in this manuscript is available online https://github.com/iabell/rapid_prototyping_COVID_19.

## References

1. National Review of Hotel Quarantine. Technical report, Australian Government Department of Health (November 2020). https://www.health.gov.au/sites/default/files/documents/2020/10/national-review-of-hotel-quarantine.pdf

2. Hellewell, J., Russell, T.W., Team, T.S.I.a.F.S., Consortium, T.C.C.-., working group, C.C.-., Beale, R., Kelly, G., Houlihan, C., Nastouli, E., Kucharski, A.J.: Estimating the effectiveness of routine asymptomatic PCR testing at different frequencies for the detection of SARS-CoV-2 infections. BMC Medicine 19(1), 106 (2021). doi:10.1186/s12916-021-01982-x. Accessed 2022-08-12

3. Brümmer, L.E., Katzenschlager, S., Gaeddert, M., Erdmann, C., Schmitz, S., Bota, M., Grilli, M., Larmann, J., Weigand, M.A., Pollock, N.R., Macé, A., Carmona, S., Ongarello, S., Sacks, J.A., Denkinger, C.M.: Accuracy of novel antigen rapid diagnostics for SARS-CoV-2: A living systematic review and meta-analysis. PLOS Medicine 18(8), 1003735 (2021). doi:10.1371/journal.pmed.1003735. Publisher: Public Library of Science. Accessed 2022-05-12

4. Dinnes, J., Deeks, J.J., Berhane, S., Taylor, M., Adriano, A., Davenport, C., Dittrich, S., Emperador, D., Takwoingi, Y., Cunningham, J., Beese, S., Domen, J., Dretzke, J., Ruffano, L.F.d., Harris, I.M., Price, M.J., Taylor-Phillips, S., Hooft, L., Leeflang, M.M., McInnes, M.D., Spijker, R., Bruel, A.V.d., Group, C.C.-.D.T.A.: Rapid, point-of-care antigen and molecular-based tests for diagnosis of SARS-CoV-2 infection. Cochrane Database of Systematic Reviews (3) (2021). doi:10.1002/14651858.CD013705.pub2. Publisher: John Wiley & Sons, Ltd. Accessed 2022-05-12

5. Forde, J.E., Ciupe, S.M.: Quantification of the Tradeoff between Test Sensitivity and Test Frequency in a COVID-19 Epidemic—A Multi-Scale Modeling Approach. Viruses 13(3), 457 (2021). doi:10.3390/v13030457. Accessed 2022-04-14

6. Steyn, N., Lustig, A., Hendy, S.C., Binny, R.N., Plank, M.J.: Effect of vaccination, border testing, and quarantine requirements on the risk of COVID-19 in New Zealand: A modelling study. Infectious Disease Modelling 7(1), 184–198 (2022). doi:10.1016/j.idm.2021.12.006. Accessed 2022-05-12

7. Garrard, G.E., Rumpff, L., Runge, M.C., Converse, S.J.: Rapid Prototyping for Decision Structuring: An Efficient Approach to Conservation Decision Analysis. In: Bunnefeld, N., Nicholson, E., Milner-Gulland, E.J. (eds.) Decision-Making in Conservation and Natural Resource Management, 1st edn., pp. 46–64. Cambridge University Press, ??? (2017). doi:10.1017/9781316135938.003.

8. Baker, C.M., Campbell, P.T., Chades, I., Dean, A.J., Hester, S.M., Holden, M.H., McCaw, J.M., McVernon, J., Moss, R., Shearer, F.M., Possingham, H.P.: From Climate Change to Pandemics: Decision Science Can Help Scientists Have Impact. Frontiers in Ecology and Evolution 10 (2022). Accessed 2022-03-29

9. Blomquist, S.M., Johnson, T.D., Smith, D.R., Call, G.P., Miller, B.N., Thurman, W.M., McFadden, J.E., Parkin, M.J., Boomer, G.S.: Structured Decision-Making and Rapid Prototyping to Plan a Management Response to an Invasive Species. Journal of Fish and Wildlife Management 1(1), 19–32 (2010). doi:10.3996/JFWM-025. Accessed 2022-10-11

10. Zelner, J., Eisenberg, M.: Rapid response modeling of SARS-CoV-2 transmission. Science 376(6593), 579–580 (2022). doi:10.1126/science.abp9498. Publisher: American Association for the Advancement of Science. Accessed 2022-05-12

11. Grabowski, F., Preibisch, G., Giziński, S., Kochańczyk, M., Lipniacki, T.: SARS-CoV-2 Variant of Concern 202012/01 Has about Twofold Replicative Advantage and Acquires Concerning Mutations. Viruses 13(3), 392 (2021). doi:10.3390/v13030392. Number: 3 Publisher: Multidisciplinary Digital Publishing Institute. Accessed 2022-10-25

12. Golding, N., Shearer, F.M., Moss, R., Dawson, P., Gibbs, L., Alisic, E., McVernon, J., Price, D.J., McCaw, J.M.: Estimating temporal variation in transmission of COVID-19 and adherence to social distancing measures in Australia, 46

13. Shearer, F.M., Moss, R., McVernon, J., Ross, J.V., McCaw, J.M.: Infectious disease pandemic planning and response: Incorporating decision analysis. PLOS Medicine 17(1), 1003018 (2020). doi:10.1371/journal.pmed.1003018. Accessed 2020-07-23

14. Duckett, S.J., Sutton, B.: On entering Australia’s third year with COVID-19. Medical Journal of Australia 215(11), 509–510 (2021). doi:10.5694/mja2.51328. Accessed 2022-12-05

