## Supplementary Material for "Rapid prototyping of models for COVID-19 outbreak detection in workplaces"

### 1 Default parameters

Here we present the default parameters used for both the exponential model (Table 1) and the agent based model (Table 2).

### 2 Defining testing schedule

For both the exponential and agent based models we define testing schedules by the number of days testing occurs per week. If testing occurs one day per week there are 7 possible testing schedules, two days per week there are 6 etc. until testing occurs 7 days per week, where there is only one possible testing schedule (see Table 3).

For the exponential model, as we increase the number of testing days, which schedule we choose becomes less important to the results (Figure 1). To calculate the detection probability, we take the average over all possible testing schedules.

For the agent based model, each simulation instance is assigned one of the possible testing schedules. To calculate the detection probability, we take the average over all simulation instances.

| Parameter name | Parameter value |
|---|---|
| $R_{eff}$ | 1.1 (a conservative estimate) |
| Test sensitivity | 85% |
| Test specificity | 100% |
| Testing days per week | 1 day |
| Generation interval | 4.7 days [**?**] [check source] |
| $I_0$ | 1 person |

**Table 1:** *Exponential model parameters*

| Parameter name | Parameter value |
|---|---|
| Duration of latent period | 1 day (transmission events are unlikely to occur on the fist day after exposure [**?**]) |
| Duration of incubation phase | Lognormal($\mu = 1.62, \sigma = 0.418$) |
| Symptomatic proportion | 2/3 of infected individuals [**?**, **?**] |
| Duration of symptomatic or asymptomatic phase | Uniformly distributed between 5 and 10 days [**?**] |
| Workplace size | 120 people |
| $R_{eff}$ | 1.1 |
| Test specificity | 100% |
| $I_0$ | 1 person |

**Table 2:** *Agent based model parameters*

| Number of testing days/week | Possible testing schedules<br>% of workforce tested<br>(Mon, Tues, Wed, Thurs, Fri, Sat, Sun) |
|---|---|
| 1 day/week | (100%, 0, 0, 0, 0, 0, 0)<br>(0, 100%, 0, 0, 0, 0, 0)<br>(0, 0, 100%, 0, 0, 0, 0)<br>(0, 0, 0, 100%, 0, 0, 0)<br>(0, 0, 0, 0, 100%, 0, 0)<br>(0, 0, 0, 0, 0, 100%, 0)<br>(0, 0, 0, 0, 0, 0, 100%) |
| 2 days/week | (100%, 100%, 0, 0, 0, 0, 0)<br>(100%, 0, 100%, 0, 0, 0, 0) … |
| ⋮ | ⋮ |
| 7 days/week | (100%, 100%, 100%, 100%, 100%, 100%, 100%) |

**Table 3:** *Possible testing schedules given number of testing days/week*

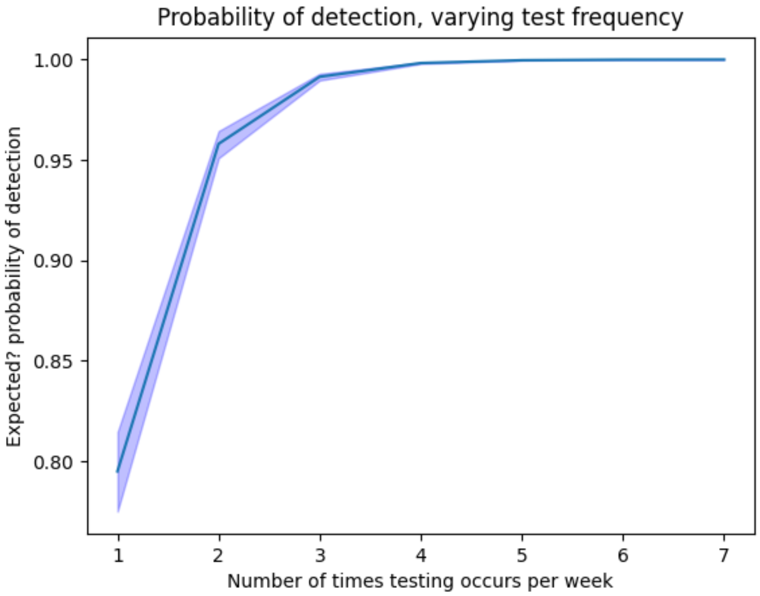


**Figure 1:** *Variance in testing schedules*
